# EUS-Guided Choledochoduodenostomy with Lumen-Apposing Metal Stent versus ERCP with Self-Expandable Metal Stent as Primary Biliary Drainage for Malignant Distal Biliary Obstruction: A Systematic Review and Meta-Analysis of Randomized Controlled Trials

**DOI:** 10.64898/2026.09.14.26363033

**Authors:** Lujing Ying, Kun He, Lisha Jing, Xi Wu, Dong Wu, Aiming Yang

## Abstract

**Background:** Whether endoscopic ultrasound-guided choledochoduodenostomy (EUS-CDS) with lumen-apposing metal stent (LAMS) is superior to endoscopic retrograde cholangiopancreatography (ERCP) with self□expandable metal stent (SEMS) as primary drainage for malignant distal biliary obstruction (MDBO) remains uncertain. We conducted a meta-analysis of randomized controlled trials (RCTs) to compare their efficacy and safety.

**Methods:** A systematic search of CENTRAL, PubMed, and Embase through September 11, 2026 was conducted. GRADE and trial sequential analysis (TSA) were performed.

**Results:** Three high-quality RCTs comprising 519 patients were included. Moderate-certainty evidence showed EUS-CDS with LAMS was associated with higher technical success (RR 1.18, 95% CI 1.09–1.28, P < 0.0001), shorter procedure time (P<0.00001), and lower postprocedural pancreatitis risk (P = 0.01) than ERCP, with comparable clinical success and overall adverse events. Sensitivity analysis, cumulative meta-analysis, and TSA supported the results.

**Conclusions:** EUS-CDS with LAMS may be considered as an alternative primary biliary drainage for MDBO.

## INTRODUCTION

Endoscopic retrograde cholangiopancreatography (ERCP) with self□expandable metal stent (SEMS) is the standard therapy for unresectable malignant distal biliary obstruction (MDBO)^1^. However, its use is limited in patients with gastric outlet obstruction (GOO) or surgically altered anatomy, and post-ERCP pancreatitis may delay subsequent anti-tumor treatment. Endoscopic ultrasound-guided biliary drainage (EUS-BD) establishes transmural biliary drainage that bypasses the duodenal papilla and was initially used as a rescue treatment after failed ERCP^2^, including EUS-guided hepaticogastrostomy (EUS-HGS) and EUS-guided choledochoduodenostomy (EUS-CDS). With improvements in devices and techniques represented by electrocautery-enhanced lumen-apposing metal stent (LAMS), the role of EUS-BD has gradually expanded. However, the efficacy and safety of EUS-CDS with LAMS versus ERCP with SEMS remain inconclusive. To systematically evaluate the performance of EUS□CDS with LAMS, we conducted a systematic review and meta□analysis of randomized controlled trials (RCTs) comparing EUS□CDS with LAMS versus ERCP with SEMS as primary biliary drainage for MDBO. To strengthen our conclusions, we used the Grading of Recommendations Assessment, Development and Evaluation (GRADE) framework and trial sequential analysis (TSA).

## METHOD

### Data sources and search strategy

We searched PubMed, Embase, and the Cochrane Central Register of Controlled Trials (CENTRAL) from inception through September 11, 2026, without language restrictions. Search terms included “EUS-CDS,” “ERCP,” “lumen-apposing metal stent,” “malignant biliary obstruction,” and “RCT,”. Full search strategies are provided in Supplementary File 1. Two reviewers (LY and LJ) independently screened studies, with disagreements resolved by discussion or a third reviewer (HK). The review followed PRISMA guidelines (Supplementary File 2) ^3^ and was registered in PROSPERO (CRD420261502602).

### Inclusion and exclusion criteria

We included RCTs comparing EUS-CDS with LAMS and ERCP with SEMS as primary biliary drainage for MDBO. Studies involving other EUS-BD approaches, rescue drainage after failed ERCP, nonrandomized studies, reviews, editorials, or insufficient data were excluded.

### Data extraction and risk of bias assessment

Two reviewers (LY and LJ) independently extracted study, patient, intervention, and outcome data. Risk of bias was assessed using the Cochrane Risk of Bias 2 (RoB 2) tool, and certainty of evidence was evaluated using the GRADE framework.

### Outcomes

The primary outcome was technical success, defined as successful completion of intended biliary drainage procedure with deployment of LAMS or SEMS. Secondary outcomes were clinical success, overall adverse events (AEs), postprocedural pancreatitis, reintervention, stent patency time, procedure time, and length of hospital stay.

### Data synthesis and statistical analysis

Pooled analyses were performed in RevMan 5.4.1 using random-effects models. Risk ratios (RRs) with 95% confidence intervals (CIs) were calculated for dichotomous outcomes and mean differences (MDs) for continuous outcomes. Medians were converted to means and standard deviations (SDs) using Wan et al.^4^. Heterogeneity was assessed using I^2^. Publication bias was not assessed because fewer than 10 studies were included. Sensitivity and cumulative meta-analysis were performed in R, and TSA was performed using TSA software (version 0.9 beta).

## RESULTS

A systematic search of three databases yielded 301 records. After removing 118 duplicates, 174 articles were excluded after reviewing the title and abstract. Of the 9 full□texts assessed, 1 was excluded because only the abstract was available, and 5 were excluded for involving techniques or stents other than EUS□CDS with LAMS (Supplementary_Figure 1). Finally, 3 RCTs comprising 519 patients (263 in the EUS□CDS group and 256 in the ERCP group)^5–7^ were included (Supplementary Table 1).

All three included RCTs had a low overall risk of bias, with uncertainty only in blinding of operators and outcome assessors (Supplementary Figure 2). GRADE assessment indicated moderate-certainty of evidence for all outcomes, detailed in Supplementary Table 2.

### Primary outcome

#### Technical success

Meta-analysis of 3 RCTs (n = 519) revealed that EUS-CDS with LAMS led to a significantly higher technical success rate than ERCP with SEMS (RR: 1.18, 95% CI: 1.09-1.28, P < 0.0001, I^2^ = 22%, moderate certainty of evidence). Leave-one-out sensitivity analysis confirmed stability of the pooled estimate (RR range: 1.15–1.22). Cumulative meta-analysis demonstrated a consistent benefit (RR=1.18, 95% CI: 1.09–1.28). TSA indicated the required information size (RIS) was 229. The cumulative sample size exceeded the RIS, confirming the robustness of this finding (Figure 1)

### Secondary outcome

#### Clinical success

Based on 3 RCTs (n = 519), meta-analysis suggested no significant difference in clinical success between the two groups for treating DMBO (RR: 1.00, 95% CI: 0.95-1.07, P = 0.88, I^2^ = 0%) (Figure 2A).

#### Overall adverse events

Meta-analysis of 3 RCTs (n = 519) showed no significant difference in overall AEs between the two groups (RR: 0.94, 95% CI: 0.68-1.30, P = 0.70, I^2^ = 0%) (Figure 2B).

#### postprocedural pancreatitis

All 3 RCTs (n = 519) recorded postprocedural pancreatitis. Meta-analysis revealed EUS-CDS with LAMS showed a significantly lower postprocedural pancreatitis rate than ERCP with SEMS (RR: 0.20, 95% CI: 0.06-0.70, P = 0.01, I^2^ = 0%) (Figure 2C).

#### Reintervention

Meta-analysis of 3 RCTs (n = 519) demonstrated no significant difference in reintervention between the two groups (RR: 0.83, 95% CI: 0.50-1.40, P = 0.49, I^2^ = 0%) (Figure 2D).

#### Stent patency time

Two RCTs reported stent patency time. Meta-analysis was not performed due to high heterogeneity (I^2^ = 72%). Although both studies showed no significant difference between the two groups, the numerical trends were inconsistent, as presented in supplementary Table 3.

#### Procedure time

The pooled analysis of 3 RCTs (n = 519) demonstrated that the EUS□CDS with LAMS had a significantly shorter procedure time compared with the ERCP with SEMS (MD = -11.60minutes, 95% CI: -14.66 - -8.54, P < 0.00001, I^2^ = 36%) (Figure 2E).

#### The length of hospital stay

Two RCTs (n = 375) reported the length of hospital stay. Meta-analysis revealed there was no significant difference between the two groups (MD = -0.32days, 95% CI: -1.78-1.14, P = 0.67, I^2^ = 0%) (Figure 2F).

## DISCUSSION

The optimal endoscopic drainage strategy for MDBO remains uncertain. Although ERCP with SEMS placement has long been the standard palliative treatment for MDBO, duodenal papilla cannulation may be technically challenging or unsuccessful in certain situations. EUS-BD offers an alternative transmural biliary drainage route, showing potential as a primary biliary drainage therapy. Our systematic review and meta-analysis of 3 RCTs including 519 patients evaluated EUS-CDS with LAMS versus ERCP with SEMS as primary biliary drainage for MDBO. Moderate-certainty evidence showed that EUS-CDS with LAMS achieved higher technical success, shorter procedure time, and lower postprocedural pancreatitis risk, with comparable clinical success and overall AEs compared to ERCP with SEMS.

The core advantage of EUS-BD is direct bile duct puncture under real-time EUS guidance without duodenal papilla cannulation, thereby overcoming limitations of ERCP in patients with GOO or difficult papillary access and potentially reducing postprocedural pancreatitis. The electrocautery-enhanced LAMS delivery system enables single-step puncture and stent placement, reducing device exchanges and shortening procedure time. Its dumbbell-shaped design provides secure anchoring between the biliary and duodenal walls, reducing stent migration risk. Moreover, EUS-CDS can often be performed without fluoroscopy, minimizing radiation exposure^6^.

Compared with previous meta-analyses^8–10^, our study showed several strengths. We included RCTs comparing EUS-CDS with LAMS versus ERCP with SEMS as first-line drainage strategy for MDBO, and selected technical success as the primary outcome, because it represents the fundamental measure of procedural feasibility for an emerging technique. In addition, we comprehensively assessed other clinically relevant outcomes including clinical success, adverse events, reintervention, stent patency, procedure time, and hospital stay. Sensitivity and cumulative meta-analyses supported the robustness of the primary outcome. Considering the limited number of eligible RCTs, TSA was performed and confirmed that the cumulative sample size exceeded the RIS for the primary outcome. Additionally, we employed the GRADE framework to assess the quality of evidence, providing more reliable evidence for clinical decision□making.

This study had some limitations. First, the total sample size remained modest despite all eligible RCTs being included. Although sensitivity analysis, cumulative meta-analysis and TSA confirmed the robustness of the primary outcome, larger-scale RCTs are needed to evaluate other important outcomes. Second, the type of SEMS used in ERCP varied across studies. Teoh et al. used partially covered SEMS^7^, while Chen et al. and Anderloni et al. used mixed types^5,6^. Additionally, Teoh et al. placed coaxial plastic stents within the LAMS during EUS-CDS^7^, which may influence stent patency assessment. Finally, cost-effectiveness and effects on subsequent chemotherapy could not be evaluated because of insufficient data and should be addressed in future RCTs.

In conclusion, moderate-certainty of evidence demonstrated EUS-CDS with LAMS was superior to ERCP with SEMS as primary biliary drainage for MDBO, with higher technical success, shorter procedure time, and comparable clinical success and overall AEs. In centers with adequate expertise, EUS-CDS with LAMS may be considered as an alternative primary biliary drainage for MDBO, particularly for patients with anticipated difficult ERCP or high risk of post□ERCP pancreatitis.

## Data Availability

All data analyzed in this study are available from the corresponding author.

## Declaration of Competing Interest

The authors declare that they have no competing interests.

## Funding

This work was supported by Natural Science Foundation of Beijing Municipality (No. J230019).

## Availability of data and materials

All data analyzed in this study are available from the corresponding author.

**Supplementary File 1:** Detailed search strategies.

**Supplementary File 2:** PRISMA guidelines.

**Supplementary File 3:** Detailed methodology.

**Supplementary Table 1:** Study characteristics.

**Supplementary Table 2:** Summary of main findings.

**Supplementary Table 3:** Stent patency time.

**Supplementary Figure 1:** The PRISMA flowchart.

**Supplementary Figure 2:** Cochrane risk-of-bias assessment.

---

**Figure 1.**
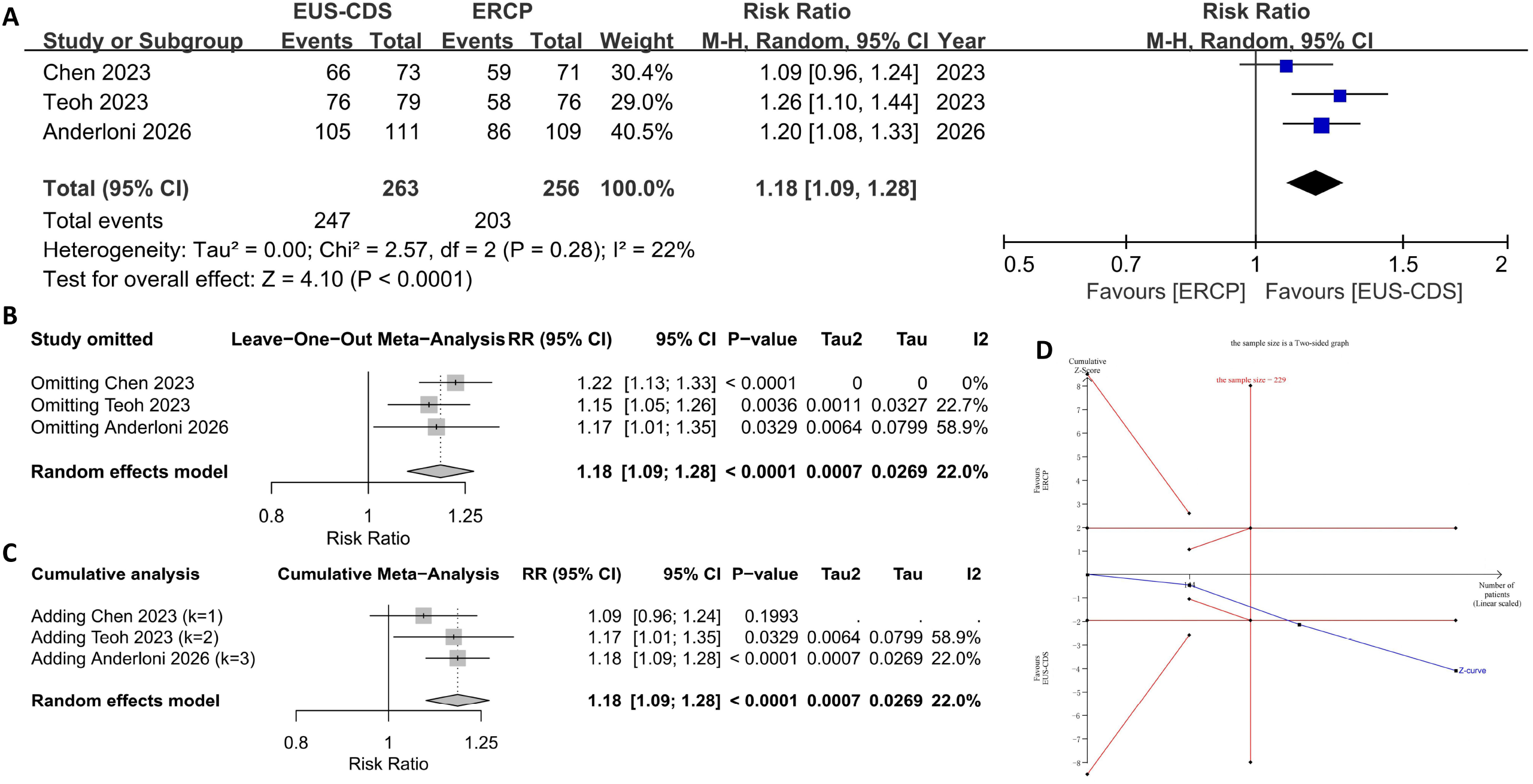
Forest plots and TSA for the primary outcome. (A) Forest plots for technical success; (B) Sensitivity analysis for technical success; (C) Cumulative analysis for technical success; (D) TSA for technical success. TSA: Trial sequential analysis; CI: Confidence interval; EUS-CDS: Endoscopic Ultrasound-Guided choledochoduodenostomy; ERCP: Endoscopic Retrograde Cholangiopancreatography.

**Figure 2.**
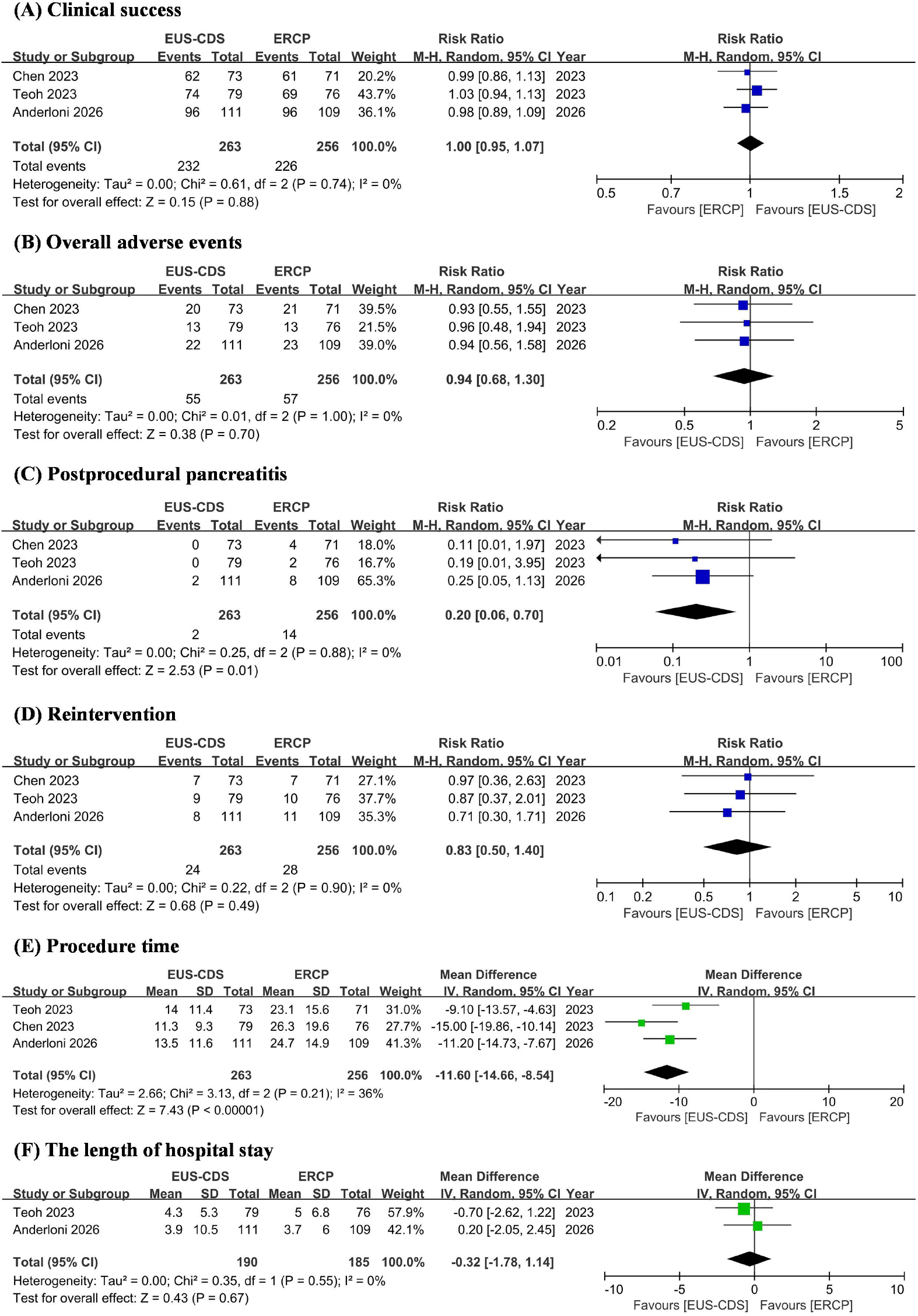
Forest plots for secondary outcomes. (A) clinical success; (B) overall adverse events; (C) postprocedural pancreatitis; (D) reintervention; (E) procedure time; (F) the length of hospital stay. CI: Confidence interval; SD: standard deviation; EUS-CDS: Endoscopic Ultrasound-Guided choledochoduodenostomy; ERCP: Endoscopic Retrograde Cholangiopancreatography.

## Notes

### Competing Interest Statement

The authors have declared no competing interest.

### Author Declarations

The study used ONLY openly available human data from the following published randomized controlled trials: Teoh AYB, et al. EUS-guided choledocho-duodenostomy using lumen apposing stent versus ERCP with covered metallic stents in patients with unresectable malignant distal biliary obstruction: A multicenter randomized controlled trial (DRA-MBO Trial). Gastroenterology. 2023;165(2):473-482. Chen YI, et al. Endoscopic ultrasound-guided biliary drainage of first intent with a lumen-apposing metal stent vs endoscopic retrograde cholangiopancreatography in malignant distal biliary obstruction: A multicenter randomized controlled study (ELEMENT Trial). Gastroenterology. 2023;165(5):1249-1261. Anderloni A, et al. Endoscopic ultrasound-guided choledochoduodenostomy vs endoscopic retrograde cholangiopancreatography in malignant distal biliary obstruction to prevent postprocedural pancreatitis: A randomized trial. Gastroenterology. 2026.

