## Supplementary Table 1 for "EUS-Guided Choledochoduodenostomy with Lumen-Apposing Metal Stent versus ERCP with Self-Expandable Metal Stent as Primary Biliary Drainage for Malignant Distal Biliary Obstruction: A Systematic Review and Meta-Analysis of Randomized Controlled Trials"

**Supplementary_Table 1.** Study characteristics

| Author, year | Intervention | Stent type | Sample size | Age (y), mean (SD) | Sex (M/F) | CBD diameter (mm), mean (SD) | Tbil (mg/dL), mean (SD) |
| --- | --- | --- | --- | --- | --- | --- | --- |
| Chen et al, 2023 | EUS-CDS | LAMS | 73 | 73.3 (10.4) | 37/26 | 17.7 (3.6) | 14.2 (7.2) |
|  | ERCP | Mixed SEMS^a^ | 71 | 70.6 (11.2) | 50/21 | 18.0 (5.0) | 16.2 (8.8) |
| Teoh et al, 2023 | EUS-CDS | LAMS | 79 | 75.1 (11.9) | 32/47 | 15.9 (3.8) | NR |
|  | ERCP | pcSEMS | 76 | 72.1 (12.4) | 41/35 | 16.8 (3.7) | NR |
| Anderloni et al, 2026 | EUS-CDS | LAMS | 111 | 73.8 (10.0) | 53/58 | 18.5 (3.5) | 13.7 (7.6) |
|  | ERCP | Mixed SEMS^a^ | 109 | 72.7 (10.1) | 47/62 | 18.0 (3.5) | 14.6 (7.6) |

EUS-CDS, Endoscopic Ultrasonography-Guided Choledochoduodenostomy; ERCP, Endoscopic Retrograde Cholangiopancreatography; LAMS, Lumen-Apposing Metal Stent; SEMS, Self-Expanding Metallic Stent; fcSEMS, fully covered SEMS; pcSEMS, partially covered SEMS; ucSEMS, uncovered SEMS; CBD, Common bile duct; TBil, Total Bilirubin; SD, standard deviation; NR, not reported.

^a^Mixed SEMS: Chen et al.: fcSEMS 47.5%, pcSEMS 13.6%, ucSEMS 39.0%; Anderloni et al.: fcSEMS and pcSEMS, proportions not reported.
