## Supplementary Table 2 for "EUS-Guided Choledochoduodenostomy with Lumen-Apposing Metal Stent versus ERCP with Self-Expandable Metal Stent as Primary Biliary Drainage for Malignant Distal Biliary Obstruction: A Systematic Review and Meta-Analysis of Randomized Controlled Trials"

**Table 2.** Summary of main findings

| Population: patients with MDBO | | | | | |
| --- | --- | --- | --- | --- | --- |
| Intervention: EUS-CDS | | | | | |
| Comparison: ERCP | | | | | |
| Outcomes | Anticipated absolute effects^*^ (95%CI) | | Relative effect (95%CI) | No. of patients (studies) | Certainty of the evidence (GRADE) |
|  | Risk with ERCP | Risk with EUS-CDS |  |  |  |
| Technical success | 793 per 1,000 | 936 per 1,000  (864 to 1000) | RR 1.18 (1.09 to 1.28) | 519 (3 RCTs) | ⊕⊕⊕◯ Moderate^a^ |
| Clinical success | 883 per 1,000 | 883 per 1,000  (839 to 945) | RR 1.00 (0.95 to 1.07) | 519 (3 RCTs) | ⊕⊕⊕◯ Moderate^a^ |
| Overall adverse events | 223 per 1,000 | 209 per 1,000  (151 to 289) | RR 0.94 (0.68 to 1.30) | 519 (3 RCTs) | ⊕⊕⊕◯ Moderate^a^ |
| Postprocedural pancreatitis | 55 per 1,000 | 11 per 1,000  (3 to 38) | RR 0.20 (0.06 to 0.70) | 519 (3 RCTs) | ⊕⊕⊕◯ Moderate^a^ |
| Reintervention | 109 per 1,000 | 91 per 1,000  (55 to 153) | RR 0.83 (0.50 to 1.40) | 519 (3 RCTs) | ⊕⊕⊕◯ Moderate^a^ |
| Procedure time | The mean procedure time of ERCP was 24.1 minutes | MD 11.6 minutes lower  (14.66 to 8.54 lower) | - | 519 (3 RCTs) | ⊕⊕⊕◯ Moderate^b^ |
| The length of hospital stay | The mean hospital stay of ERCP was 4.2 days | MD 0.32 days lower  (1.78 lower to 1.14 higher) | - | 375 (2 RCTs) | ⊕⊕⊕◯ Moderate^a^ |

GRADE Working Group grades of evidence. High Certainty: further research is very unlikely to change our confidence in the estimate of effect; Moderate Certainty: further research is likely to have an important impact on our confidence in the estimate of effect and may change the estimate; Low Certainty: further research is very likely to have an important impact on our confidence in the estimate of effect and is likely to change the estimate; Very low quality: we are very uncertain about the estimate; CI: confidence interval; RR: Relative risk; EUS-CDS: endoscopic ultrasound-guided choledochoduodenostomy; ERCP: endoscopic retrograde cholangiopancreatography. ^*^The basis for the assumed risk is the average control group proportion across all comparisons. ^a^downgraded one level for imprecision, ^b^downgraded one level for inconsistency.
