## Supplementary Table 3 for "EUS-Guided Choledochoduodenostomy with Lumen-Apposing Metal Stent versus ERCP with Self-Expandable Metal Stent as Primary Biliary Drainage for Malignant Distal Biliary Obstruction: A Systematic Review and Meta-Analysis of Randomized Controlled Trials"

**Supplementary Table 3.** Stent patency time (days)

| **Author, year** | **EUS-CDS** | | **N** | **ERCP** | | **N** | **P value** |
| --- | --- | --- | --- | --- | --- | --- | --- |
| Chen, 2023 | Mean: 163.9 | SD: 128.4 | 73 | Mean: 200.1 | SD: 135.5 | 71 | 0.10 |
| Teoh, 2023 | Mean: 183.2 | SD: 131.3 | 79 | Mean: 161.3 | SD: 136.0 | 76 | 0.81 |

EUS-CDS: endoscopic ultrasound-guided choledochoduodenostomy; ERCP: endoscopic retrograde cholangiopancreatography; SD: standard deviation.
