## Supplementary File 1 for "EUS-Guided Choledochoduodenostomy with Lumen-Apposing Metal Stent versus ERCP with Self-Expandable Metal Stent as Primary Biliary Drainage for Malignant Distal Biliary Obstruction: A Systematic Review and Meta-Analysis of Randomized Controlled Trials"

**PUBMED**

| No. | Query | Results |
|---|---|---|
| #1 | "EUS"[Title/Abstract] OR "EUS-guided"[Title/Abstract] OR "endoscopic ultrasound"[Title/Abstract] OR "endosonography"[MeSH Terms] | 29,759 |
| #2 | "biliary drainage"[Title/Abstract] OR "choledochoduodenostomy"[Title/Abstract] | 7,183 |
| #3 | "cholangiopancreatography, endoscopic retrograde"[MeSH Terms] OR "ERCP"[Title/Abstract] | 26,278 |
| #4 | biliary obstruction[Title/Abstract] | 6,749 |
| #5 | "Randomized Controlled Trial"[Publication Type] OR "Controlled Clinical Trial" [Publication Type] OR "Randomized Controlled Trial"[Title/Abstract] OR "Controlled Clinical Trial"[Title/Abstract] OR "randomized"[Title/Abstract] OR "randomised"[Title/Abstract] OR "randomly"[Title/Abstract] | 1,583,498 |
| #6 | #1 AND #2 AND #3 AND #4 AND #5 | 60 |

**EMBASE**

| No. | Query | Results |
|---|---|---|
| #1 | 'endoscopic ultrasonography'/exp OR eus:ab,ti OR 'eus guided':ab,ti OR 'endoscopic ultrasound guided':ab,ti | 65,451 |
| #2 | 'biliary tract drainage'/exp OR 'biliary drainage':ab,ti OR choledochoduodenostomy:ab,ti | 39,369 |
| #3 | #1 AND #2 | 4,870 |
| #4 | 'endoscopic retrograde cholangiopancreatography'/exp OR 'endoscopic retrograde cholangiopancreatography':ab,ti OR ercp:ab,ti | 63,577 |
| #5 | 'biliary obstruction':ab,ti | 11,983 |
| #6 | 'randomized controlled trial'/exp OR 'controlled clinical trial'/exp OR 'randomized controlled trial':ab,ti OR 'controlled clinical trial':ab,ti OR randomized:ab,ti OR randomised:ab,ti OR randomly:ab,ti | 2,511,997 |
| #7 | #3 AND #4 AND #5 AND #6 | 161 |

**CENTRAL**

| No. | Query | Results |
|---|---|---|
| #1 | MeSH descriptor: [Endosonography] explode all trees | 591 |
| #2 | (EUS):ti,ab,kw OR (EUS-guided):ti,ab,kw OR (endoscopic ultrasound):ti,ab,kw | 2883 |
| #3 | #1 OR #2 | 3135 |
| #4 | (biliary drainage):ti,ab,kw OR (choledochoduodenostomy):ti,ab,kw | 1097 |
| #5 | #3 AND #4 | 206 |
| #6 | (endoscopic retrograde cholangiopancreatography):ti,ab,kw OR (ERCP):ti,ab,kw | 3303 |
| #7 | (biliary obstruction):ti,ab,kw | 991 |
| #8 | MeSH descriptor: [Randomized Controlled Trial] explode all trees | 34 |
| #9 | MeSH descriptor: [Controlled Clinical Trial] explode all trees | 37 |
| #10 | (randomized controlled trial):ti,ab,kw OR (controlled clinical trial):ti,ab,kw OR (randomized):ti,ab,kw OR (randomised):ti,ab,kw OR (randomly):ti,ab,kw | 1507054 |
| #11 | #8 OR #9 OR #10 | 1507054 |
| #12 | #5 AND #6 AND #7 AND #11 | 80 |
