## Supplementary figures and images for "EUS-Guided Choledochoduodenostomy with Lumen-Apposing Metal Stent versus ERCP with Self-Expandable Metal Stent as Primary Biliary Drainage for Malignant Distal Biliary Obstruction: A Systematic Review and Meta-Analysis of Randomized Controlled Trials"

### Supplementary Figure 1

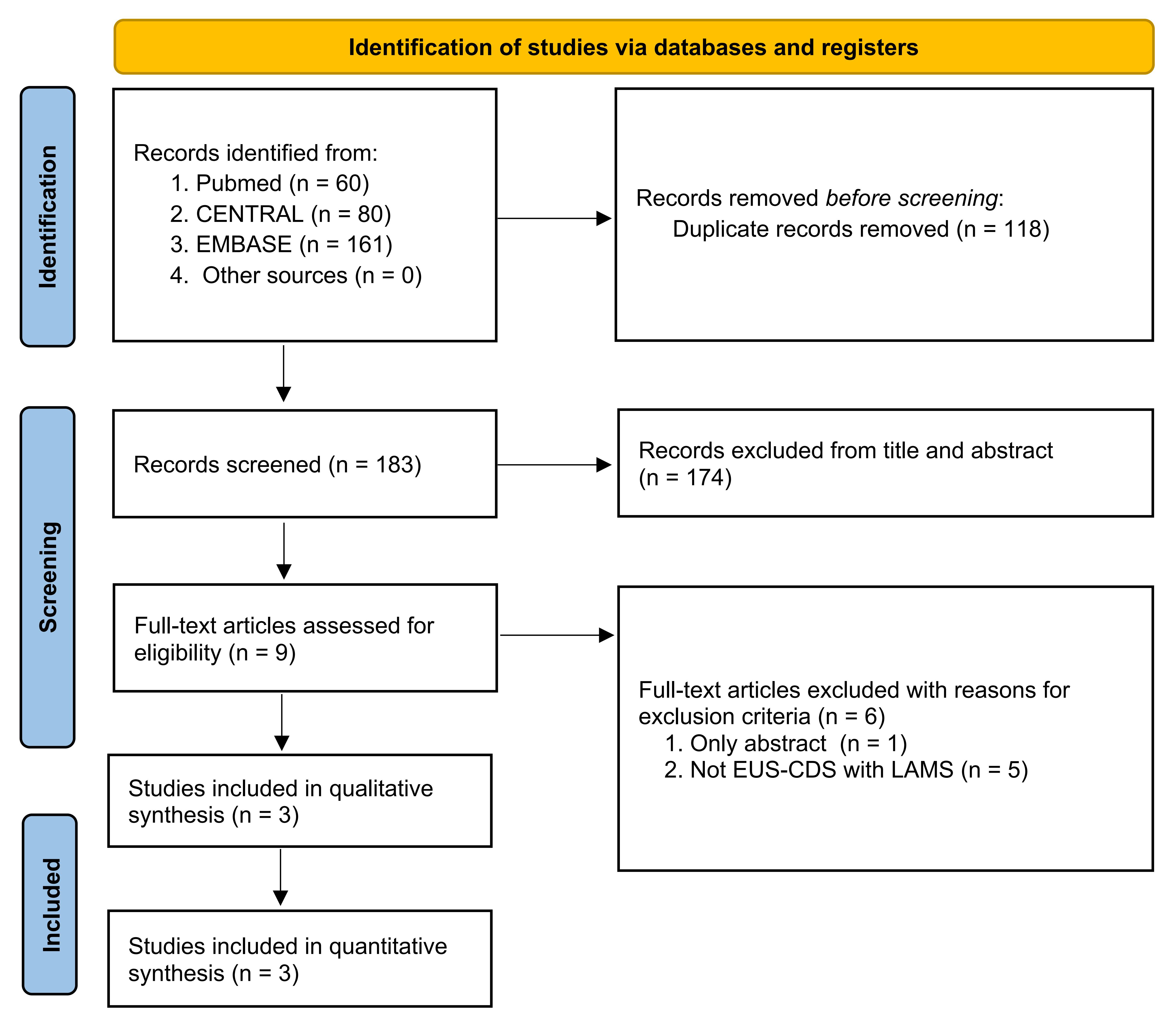

### Supplementary Figure 2

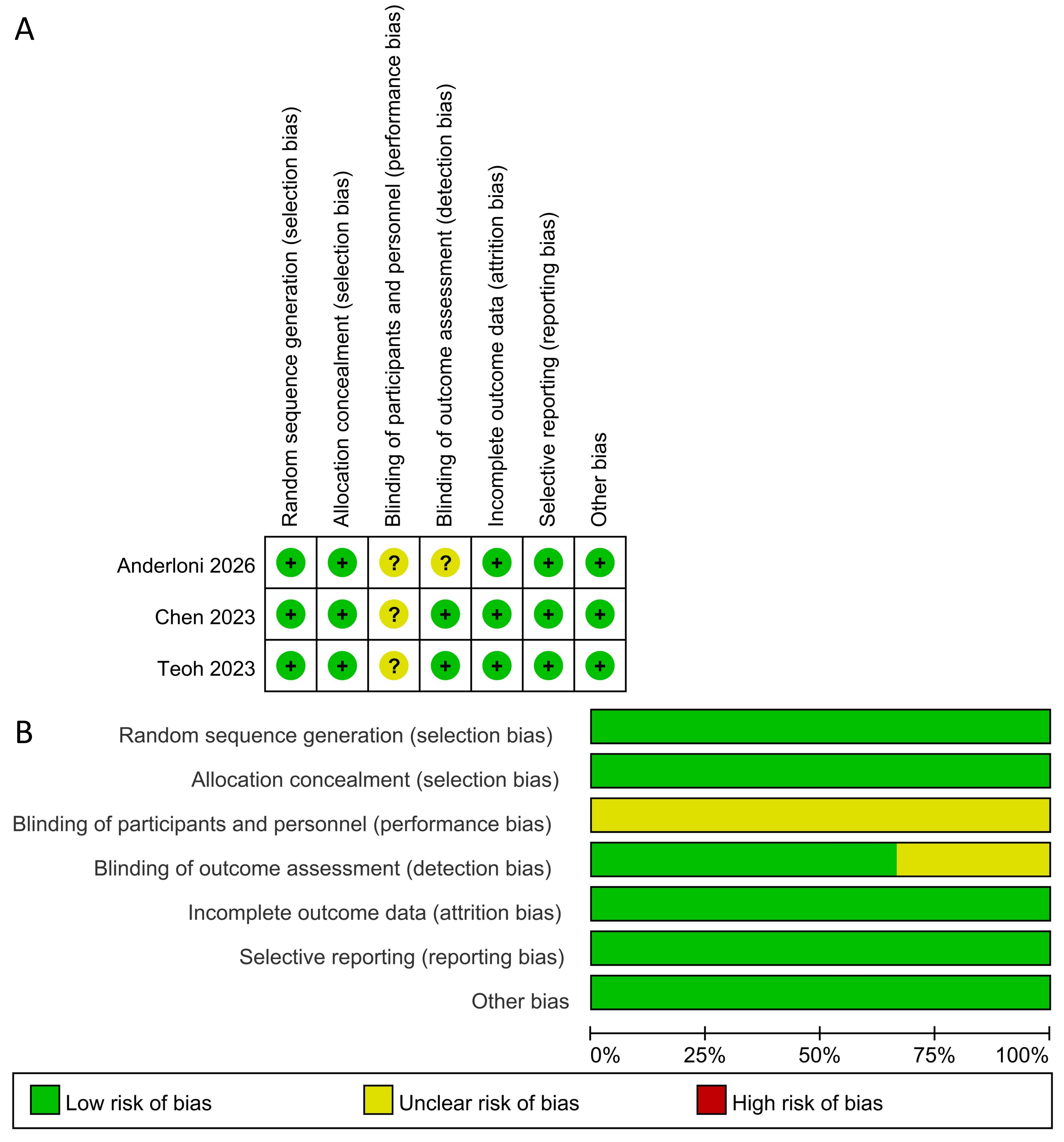
